# Key performance indicators of COVID-19 contact tracing in Belgium from September 2020 to December 2021

**DOI:** 10.1101/2022.10.04.22280542

**Authors:** Cécile Kremer, Lander Willem, Jorden Boone, Wouter Arrazola de Oñate, Naïma Hammami, Christel Faes, Niel Hens

## Abstract

**Background:** Contact tracing aims to prevent onward transmission of infectious diseases and data obtained during tracing provide unique information on transmission characteristics. A key performance indicator that has been proposed to evaluate contact tracing is the proportion of cases arising from known contacts. However, few empirical studies have investigated the effectiveness of contact tracing.

**Methods:** Using data collected between September 2020 and December 2021 in Belgium, we investigated the impact of contact tracing on SARS-CoV-2 transmission. We compared confirmed cases that were previously identified as a close contact to those that were not yet known, in terms of their traced contacts and secondary cases as well as the serial interval. In addition, we established contact and transmission patterns by age.

**Findings:** Previously traced, hence ‘known’, cases comprised 20% of all cases and they were linked to relatively fewer close contacts as well as fewer secondary cases and a lower secondary attack rate compared to cases that were not already known. In addition we observed a shorter serial interval for ‘known’ cases. There was a relative increase in transmission from children to adults during circulation of the Delta and Omicron variants, without an increase in the extent of contact between these age groups.

**Interpretation:** These results suggest that contact tracing in Belgium has been effective in reducing onward transmission and that individuals aware of their exposure to SARS-CoV-2 seemed more reserved in their social contact behaviour. Data from a reference period or region are needed to measure the impact of contact tracing in terms of the number of cases and deaths averted.

## Introduction

Coronavirus disease 2019 (COVID-19), caused by the severe acute respiratory syndrome coronavirus 2 (SARS-CoV-2), emerged in December 2019 and led to a global cumulative number of confirmed cases of over 598 million and a cumulative number of reported deaths over 6·4 million by the end of August 2022.^1^ In order to maintain the healthcare system, non-pharmaceutical interventions (NPIs) have been adopted during the pandemic in many countries. In the initial phase of the pandemic, many countries imposed a lockdown to restrict all social gatherings. When restrictions were relieved, countries implemented contact tracing in the philosophy of having person-tailored quarantine measures instead of nationwide measures. Active case finding and swift contact tracing have been effective for tuberculosis prevention and early treatment for many years.^2,3^ However, the required scale for COVID-19 was unseen in many countries. Effective contact tracing can, when carried out quickly, prevent onward transmission from newly infected individuals and their possibly infected contacts. Contact tracing also enhances active surveillance and provides unique data for estimating transmission characteristics such as secondary attack rates, risk factors associated with infection, and important parameters determining the speed at which a virus is spreading. Timely notification of potential exposure to SARS-CoV-2 is essential to break transmission chains, as indicated by several mathematical modelling studies investigating the impact of contact tracing.^4–8^ A simulation-based study for Belgium concluded that an average reduction in hospital admissions between 22-57% is possible with contact tracing in place, when assuming that 70% of symptomatic cases are subjected to contact tracing and comply with home isolation.^4^ A recent systematic review reported that provider-initiated contact tracing was generally associated with improved control of communicable diseases, while also identifying the need for quantitative studies on the effectiveness of contact tracing.^9^

It is not trivial to quantify the effectiveness of contact tracing in the absence of a reference period or region in which the same data are collected without informing risk contacts of their potential exposure and the importance of quarantining. In England, a technical error in the contact tracing system in late September 2020 resulted in a failure to provide timely contact tracing for around 20% of all cases at that time. It was later found that cases that were traced according to the standards in place were associated with a 63% reduction in secondary infections across the six weeks following this error, compared to those cases that were accidentally missed by the tracing system.^10^ One of the key performance indicators (KPIs) for contact tracing proposed by the ECDC and WHO is the proportion of new cases arising from known contacts, giving an indication of the quality and completeness of the contact tracing system by pointing out the capacity of the system to identify all potential cases.^11,12^ As such, a higher proportion of cases that were previously identified as a contact indicates that more individuals at risk of infection are reached by contact tracing. Other indicators include delay distributions such as the time between the clinical test and contacting the contacts of an index case.

In this study, we used contact tracing data from Flanders and Brussels-Capital region, Belgium, collected between September 2020 and December 2021. During this period, several variants of concern (VoC) have been circulating in Belgium. In December 2020, there was no circulation of a VoC yet and the contact tracing system had recovered from the severe second COVID-19 wave in Belgium. In April and July 2021, there was circulation of the Alpha and Delta VoC, respectively. The Omicron VoC started circulating in Belgium in December 2021, though it had not yet taken over dominance from the Delta VoC.^13^ Contact tracing in Belgium was implemented in May 2020 by the regional authorities (Flanders, Brussels-Capital region, and Wallonia) and coincided with the deconfinement of several restrictive measures from the initial lockdown. The initial phase of the contact tracing encountered issues due to a changing test strategy and limited capacity in combination with technical aspects. Therefore, we started our analysis after the Belgian summer holidays, in September 2020, when the system was stable and up for four months. We investigated several key indicators and discuss their opportunities and pitfalls. In particular, we explored (1) the proportion of all SARS-CoV-2 confirmed cases that were previously traced as a high- or low-risk contact, i.e. ‘known’ cases, (2) the number of traced contacts for ‘known’ and ‘new’ cases, and (3) the number of secondary cases for ‘known’ and ‘new’ cases. In addition, we investigated contact and transmission patterns based on these data.

## Methodology

### Description of the contact tracing system

On 7 May 2020, the Flemish Agency for Care and Health (‘Vlaams Agenstschap Zorg en Gezondheid’) started with contact tracing in order to identify risk contacts of SARS-CoV-2 confirmed cases and alert them of possible exposure to the virus. Initially, the system operated using ‘batches’ of index cases and their contacts, making it difficult to distinguish between delays due to slow tracing or technical delays. In September 2020, the system was implemented in real-time. By the end of December 2021 the majority of the Belgian population was fully vaccinated, hence testing and NPIs were gradually curtailed. Therefore, our analysis is based on the data collected between September 2020 and December 2021. An index case was defined as a confirmed case whose detection initiated a contact tracing event. Index cases were queried about any contacts that occurred from within the two days before symptom onset, or within the two days before their positive SARS-CoV-2 test result in case of no symptoms. Inquiries were done via telephone by call agents at centralised locations. Contacts reported by index cases were categorized as high- or low-risk. High-risk contacts (HRC) were defined as either physical contacts, or non-physical contacts with a duration of more than 15 minutes within 1·5m distance without correct use of face masks. Other contacts not meeting the conditions for high-risk were deemed low-risk contacts (e.g. non-physical contact within 1·5m distance with correct use of face masks).

Interview data from confirmed COVID-19 cases who were contacted as an index case contained demographic information, the number of reported contacts (i.e. the number of unique persons an index case had met in the last 48 hours before their positive test or symptom onset), the number of registered contacts (i.e. the number of reported contacts that were registered in the contact tracing database), and information on the traced contacts (i.e. registered contacts that were effectively reached by the contact tracing). The number of traced contacts was lower than the number of reported/registered contacts because some contacts could not be reached or due to incomplete contact information. In addition, contacts that were reported by more than one index case were usually only traced once. Household members of a confirmed case that had already been quarantined were also not always traced individually. Interview data were also available from low- and high-risk contacts of index cases, and we included those for which a national registry number (NRN) was available such that they could be identified as an index case after testing positive. Contacts that were linked to the same index case more than once or that could not be linked to an index case due to missing information (e.g. when the reported index case is not a resident of the included regions) were excluded from the analyses. In collectivities such as hospitals, schools, and nursing homes only individual contacts occurring outside these settings were available. Contacts occurring within a collectivity were followed up by the responsible medical services and were only available in aggregated format. All data were encrypted by the Flemish Agency for Care and Health before being made available for analysis.

### Key performance indicators

One of the ECDC’s key performance indicators (KPIs) for contact tracing is the proportion of new cases arising from known contacts.^11,12^ These cases have previously received information regarding their potential exposure to the virus, as well as information on testing and quarantine, and we defined them as ‘known’ index cases. In contrast, we defined ‘new’ index cases as individuals that tested positive for SARS-CoV-2 but were not previously identified as a high- or low-risk contact. We investigated differences between these ‘new’ and ‘known’ index cases in terms of the number of traced contacts and secondary cases, in order to describe the impact of contact tracing on onward transmission. Due to the lack of reliable household information for the entire study period from September 2020 to December 2021, initially we did not make a distinction between household and non-household contacts. Since the end of May 2021, the identifier for traced HRC included an indicator for household status. Therefore, for the period from June 2021 to December 2021, we can divide HRC into household and non-household contacts. Delay distributions were constructed to investigate how quickly contact tracing was carried out. The time between symptom onset of an index case and their SARS-CoV-2 test, as well as between symptom onset and receiving the result of that test, give an indication of how quickly individuals get tested after showing symptoms and how quickly the test results are available. On the other hand, the time between the index case receiving their positive test result and getting contacted by the tracing system gives an indication of how quickly the contact tracing system operates.

### Contact and transmission patterns

Based on the available data sources we constructed a contact line list in which each traced contact with known NRN was linked to their index case. A transmission line list was constructed by linking infected contacts to their reported index case if a positive test occurred in the 21 days before or after their index case tested positive.^14^ If not, transmission between these individuals was deemed very unlikely and the pair was only kept in the contact line list. For non-positive contacts, we had no information on whether they tested negative or were not tested. Sequencing information to identify VoCs was available for a limited number of cases. When sequencing information was available for both individuals in a potential transmission pair, but different VoCs were identified, this pair was removed from the transmission line list. We constructed age-specific contact and transmission matrices for specific months to investigate the dynamics for different SARS-CoV-2 VoCs. We assumed transmission would have occurred from index to contact. The contact matrices represent the number of contacts between an index in age group *i* and a contact in age group *j*, divided by the total number of index cases in age group *i*. The transmission matrices represent the number of infections in age group *j* that can be linked to an index case in age group *i*, divided by the total number of infections in age group *j*. Another important transmission characteristic is the serial interval, defined as the time between symptom onset in the index case and symptom onset in their secondary case, which gives an indication of the speed of transmission. We calculated the serial interval over time as the monthly moving average by date of the positive test of the index case (i.e. forward serial interval), assuming that the index case was the source of infection and randomly selecting one index case when there were multiple possibilities. The serial interval was constrained to lie within the biologically plausible interval of -5 to 21 days.^15^

Negative binomial regression models were used to investigate the relationship between the number of traced (positive) HRC and age over time. Age and time were modelled using natural cubic splines, with degrees of freedom selected based on Akaike Information Criterion (AIC).^16^ A training sample comprising 70% of all observations was used to determine the best model, and then validated based on the fit to the remaining observations. For these analyses, due to sparseness of the data for index cases aged above 85 years, we included index cases until age 85 (comprising 98% of all index cases) for the period from December 2020 to December 2021.

## Results

The data as extracted on 25 May 2022 included 1 004 694 index cases (987 210 unique individuals) identified between September 2020 and December 2021. During this period, 1 092 985 traced contacts (917 102 unique individuals) could be linked to 416 645 unique index cases (42·2% of all available index cases). Figure 1 shows the number of index cases by age group, together with the most important imposed control measures regarding social contacts. In the remainder of this work we will focus on HRC since these make up the majority of traced contacts (94·9%). Among index cases for whom at least one HRC was registered in the tracing system, the average number of registered HRC was 2·8 (IQR 1 - 4). Overall, 76·4% of registered HRC were effectively traced (Figure S1). Among index cases linked to at least one traced HRC, the average number of traced HRC was 2·5 (IQR 1 - 3). During the period from June 2021 to December 2021 (which includes 51·1% of all available index cases), 46·6% of all traced HRC were defined as household contacts.

**Figure 1:**
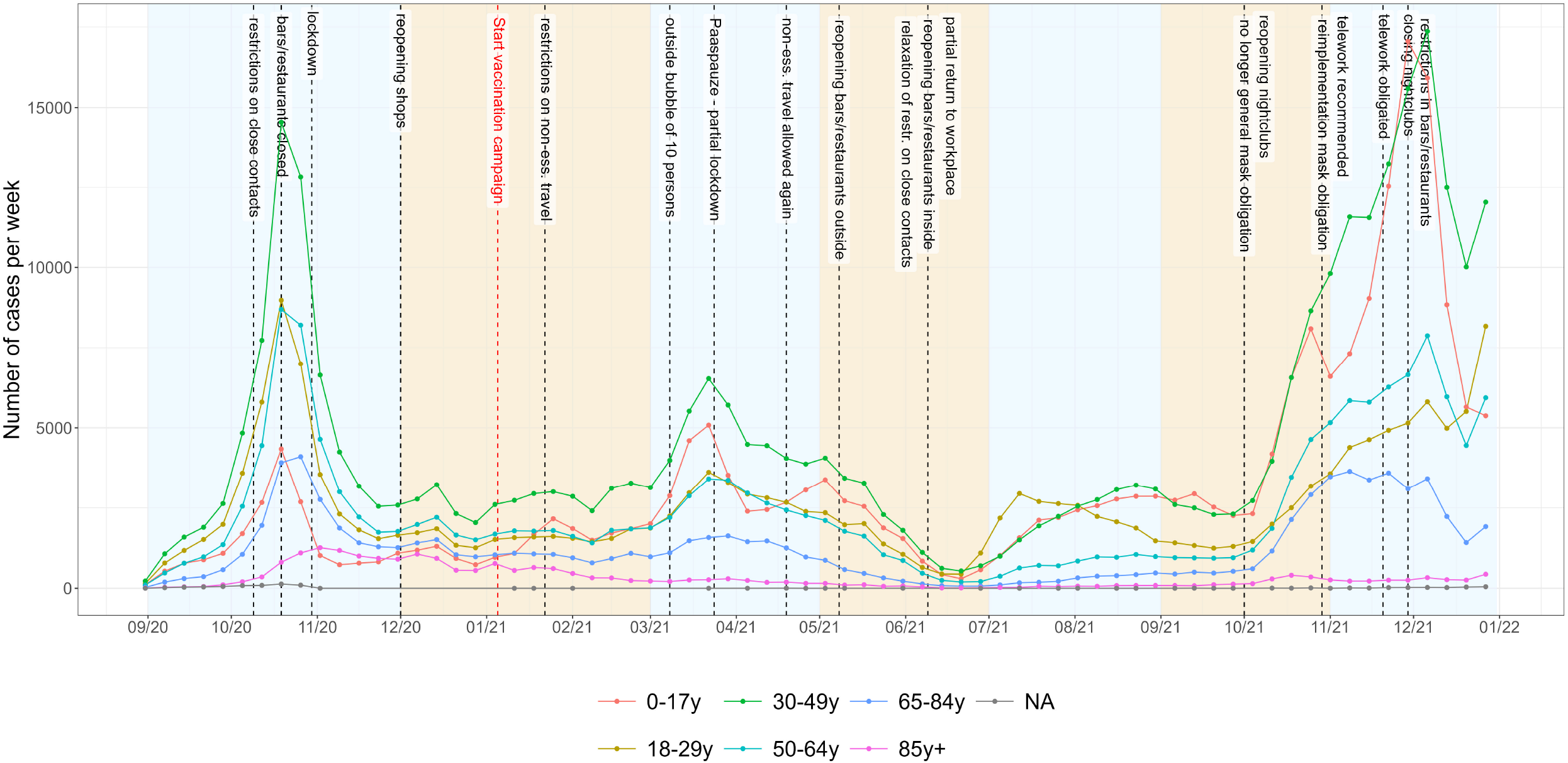
Number of index cases that were contacted during the period from September 2020 to December 2021 by age group, with an overview of the most influential control measures regarding social contacts.

### Key performance indicators

During the investigated period from September 2020 to December 2021, 19·1% of all index cases had been previously identified as a risk contact (i.e. ‘known’ index cases), of which 96·7% as HRC. Starting at less than 10% during September–November 2020, the proportion of ‘known’ index cases increased to around 25% during May–June 2021 (Figure 2a). The proportion of ‘known’ index cases was highest among 0-17 year olds, and lowest among those above 65 years old. Changes in the proportion of ‘known’ index cases over time followed a similar trend within each age group (Figure S2). Index cases that were not previously identified as a risk contact (i.e. ‘new’ index cases) were linked to a higher average number of traced HRC compared to ‘known’ index cases (Figure 2b). For both ‘known’ and ‘new’ index cases we observed an increasing trend in the number of traced HRC, which may be explained by the relaxation of control measures over time. A drop in the number of traced HRC is observed during November–December 2021, with no substantial difference between ‘known’ and ‘new’ index cases during that period. In general, the average number of traced HRC was higher for ‘new’ index cases regardless of household status, though the difference with ‘known’ index cases was less pronounced for household HRC (Figure S3a).

**Figure 2:**
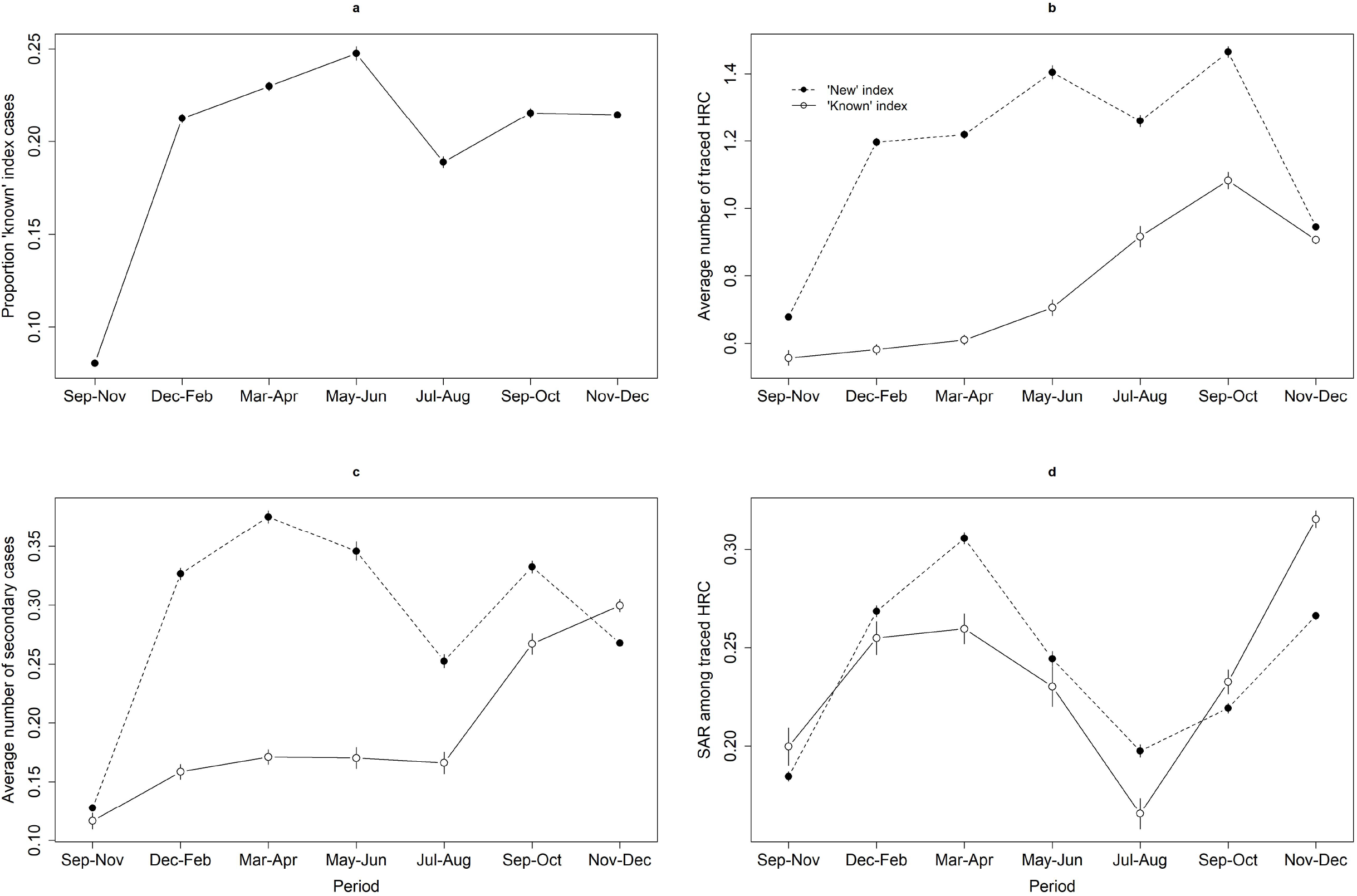
Evolution in the (a) proportion of index cases that were previously identified as a risk contact, (b) average number of traced high-risk contacts (HRC) for ‘new’ and ‘known’ index cases, (c) average number of secondary cases for ‘new’ and ‘known’ index cases, and (d) secondary attack rate (SAR) among traced HRC of ‘new’ and ‘known’ index cases. Vertical bars represent the mean *±* 2*×* standard error.

The average number of traced secondary cases was relatively stable for ‘known’ index cases until September 2021, and lower compared to the number of secondary cases linked to ‘new’ index cases (Figure 2c). In addition to the absolute number of secondary cases, we also compared the secondary attack rate (SAR), representing the number of secondary cases among all contacts. For contacts that could be linked to multiple index cases (13·7% of all HRC), we randomly selected one index case when calculating the SAR. The SAR was generally lower for ‘known’ index cases compared to ‘new’ index cases, until September 2021 (Figure 2d). While the number of traced household HRC remained relatively stable over time, an increase was observed in the number of secondary cases and SAR among household contacts (Figure S3c).

In the fall of 2020, Belgium experienced a severe second COVID-19 wave during which test capacity was under great pressure, resulting in a temporary change in testing strategy. The burden on contact tracing also increased, as reflected by a sharp increase in the average number of calls per agent (Figure S4). As a result, only index cases were called during that time and their HRC were informed via text message. More specifically, between 16 October and 2 November 2020, text messages were sent to most HRC, while low-risk contacts were not traced at all. As a consequence, it is not possible to link index cases and their contacts and to identify ‘known’ index cases during September–November 2020. This artefact disturbs the indicators we used since these assume a standard data flow. Of the 189 900 index cases contacted between September and November 2020, 8·1% had been previously identified as a risk contact. This proportion increased between 1 September and 12 October, after which it decreased until its lowest point around 31 October 2020 before increasing again. During the period around 31 October 2020, there was no substantial difference in the number of traced HRC and secondary cases between ‘known’ and ‘new’ index cases, while it was observed that ‘known’ index cases were associated with fewer HRC and secondary cases for the remaining period (Figure S5).

For symptomatic index cases (data available from November 2020 onward), the average time between symptom onset and taking a SARS-CoV-2 test was 2·3 days, with 90% of index cases getting tested within 5 days after symptom onset. The average time between symptom onset and their positive test result was 2·9 days, with 90% of index cases receiving their test results within 6 days after symptom onset. It should be noted that 3·7% of symptomatic index cases got tested before showing symptoms. The average time between receiving the positive test result and being contacted by the tracing system was 1·3 days, with 90% of index cases being contacted within 3 days of their test result (Figure 3).

**Figure 3:**
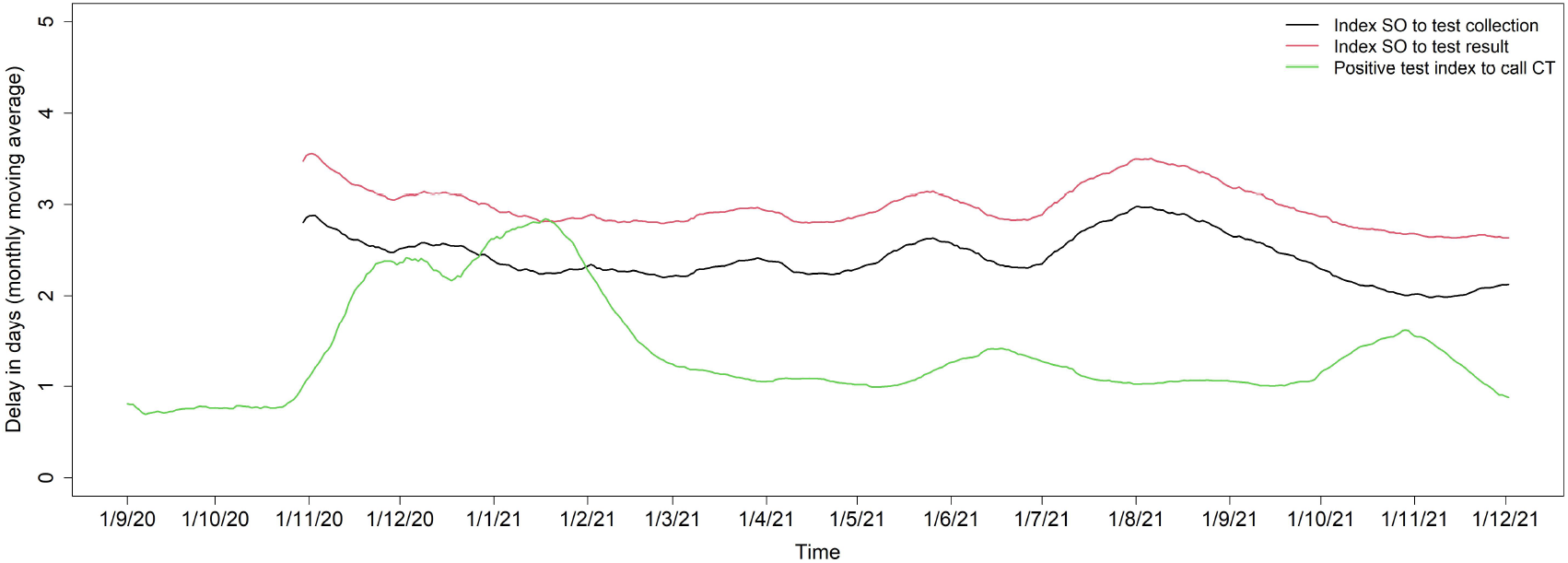
Delay distribution for the time between index symptom onset and test collection (black), between index symptom onset and test result (red), and between positive test index and the call from the tracing center (green).

### Contact and transmission patterns

Figure 4 shows the age-specific contact and transmission matrices for December 2020, April 2021, July 2021, and December 2021. During each period, we observed an assortative relation in both the contact and transmission matrices, i.e. most contacts and transmission occurred within age groups. In July 2021, the average number of contacts between individuals aged 20-29 years was highest, which is also reflected in the transmission matrices. An increase in transmission from 10-19 year olds to 40-49 year olds is observed during July 2021 (i.e. Delta VoC), while the average number of traced contacts between these age groups remained relatively stable over time. Similarly, in December 2021 (i.e. rise of the Omicron VoC), we observe an increase in transmission from 0-9 year olds to 30-39 year olds. The serial interval was shortened over time, in line with previous studies.^17,18^ In addition, we found that the serial interval was shorter for ‘known’ index cases compared to ‘new’ index cases from July 2021 onward (Figure 5).

**Figure 4:**
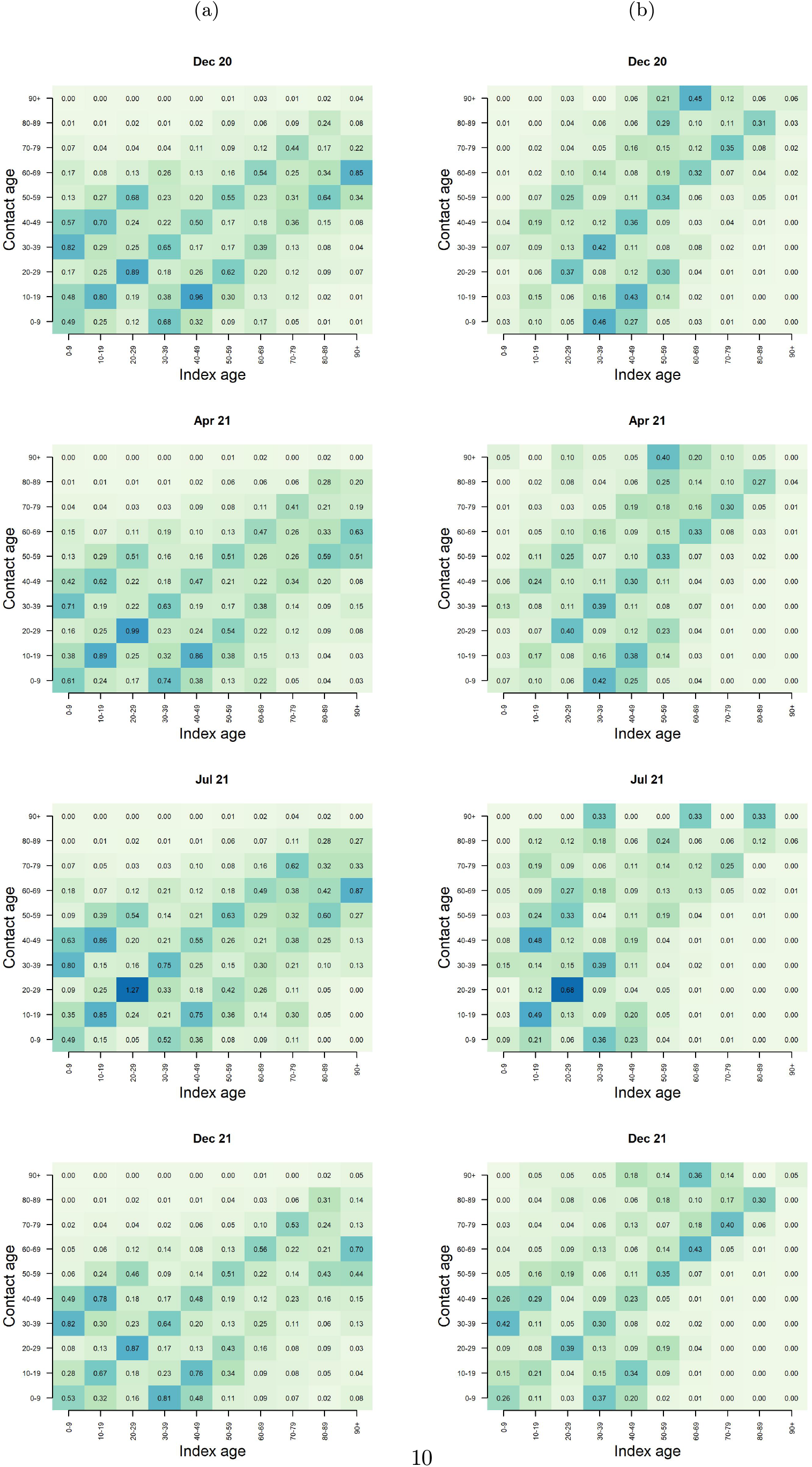
Age-specific (a) contact and (b) transmission matrix for December 2020, April 2021 (i.e. Alpha VOC), July 2021 (i.e. Delta VOC), and December 2021 (i.e. Omicron VOC).

**Figure 5:**
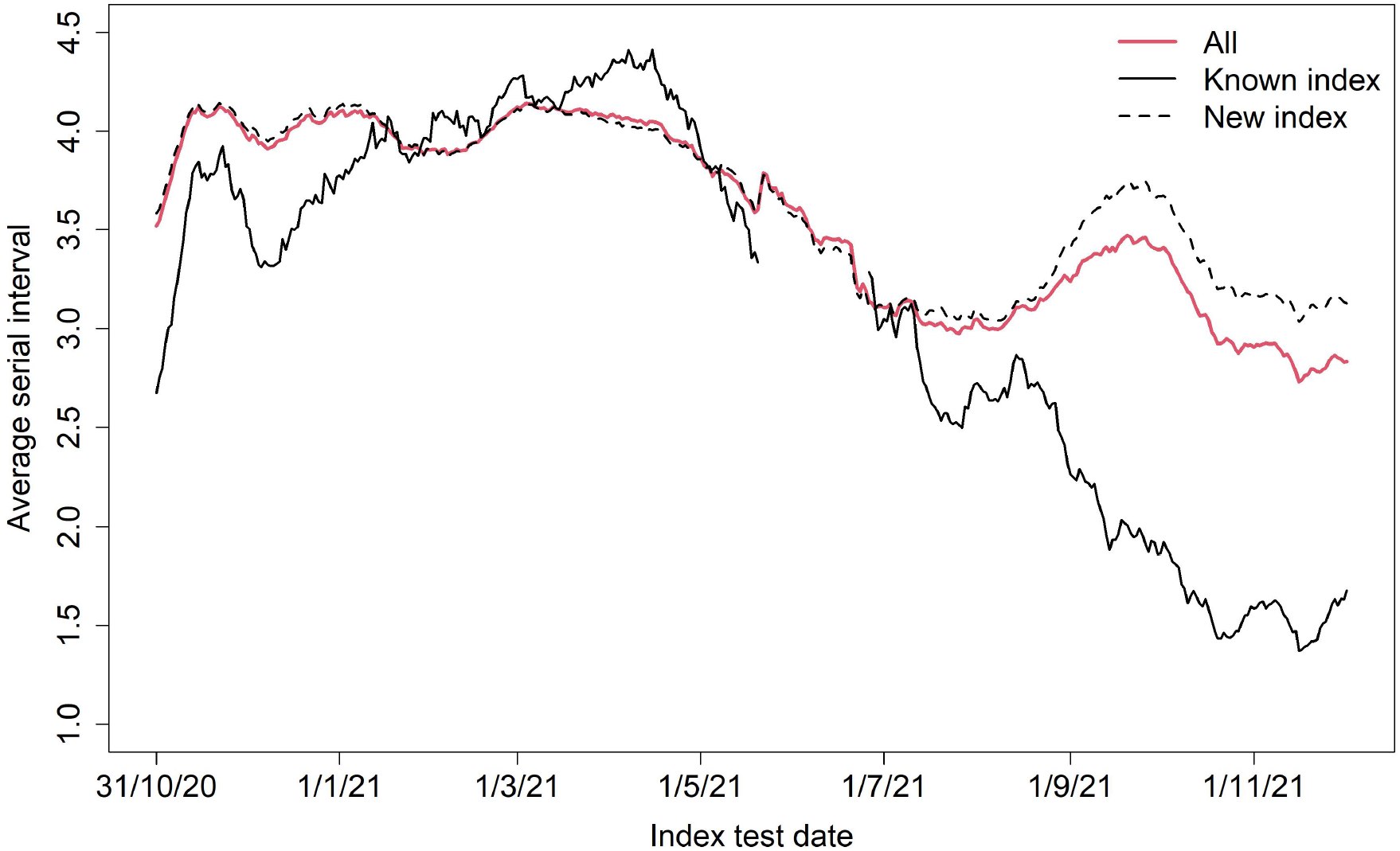
Monthly moving average empirical serial interval (in days; 116 453 transmission pairs included; 15 190 transmission pairs with ‘known’ index and 101 263 with ‘new’ index included).

For the period from December 2020 to December 2021, we investigated the influence of age of the index case on the number of traced HRC and secondary cases over time using negative binomial regression models. Initially, many traced HRC were linked to index cases aged around 10, 20, and 40 years (Figure 6a), with relatively more secondary cases linked to index cases aged around 40 (Figure 6b). Around March 2021, we observe an increase in the number of secondary cases linked to index cases aged below 10 years old (Figure 6b), without an increase in the number of traced HRC (Figure 6a). A similar trend, though not as obvious, is observed in April 2021 in Figure 4.

**Figure 6:**
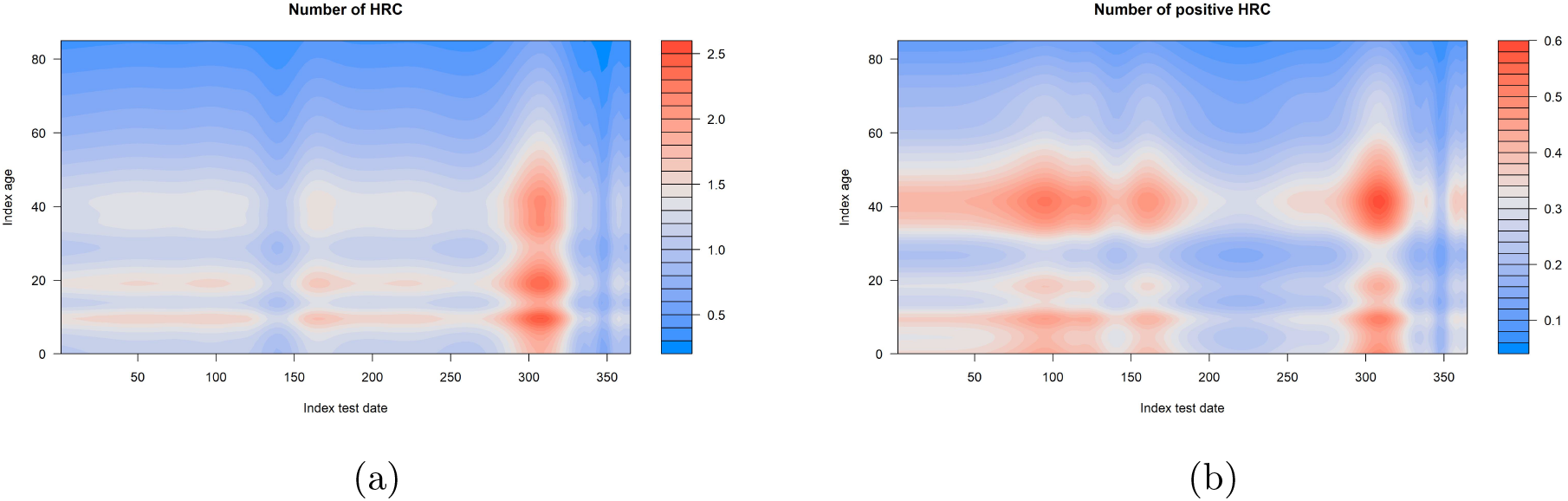
Influence of test result date and age of the index case on the number of (a) traced HRC and (b) secondary cases. Index test dates start at 1 December 2020.

## Discussion

In this study, we found that SARS-CoV-2 confirmed cases that had previously been identified as a risk contact generally reported fewer HRC and were linked to fewer secondary infections, as well as a lower attack rate among their HRC. These results suggest that individuals that were informed about their exposure to an infected person were more reserved in their social contact behaviour, also reflecting the obligated quarantine for HRC. KPIs obtained during periods of high burden on the healthcare system should be interpreted with caution due to possible changes in testing and quarantine strategies. For example during the period September–November 2020, we observed a decrease in the proportion of index cases that were previously identified as a risk contact, coinciding with a strong peak in SARS-CoV-2 incidence in the population resulting in changes in testing and tracing strategies. After the summer of 2021, the contact tracing strategy changed to an approach where index cases who had attended some sort of gathering were queried in more detail when there was at least one other index linked to the same gathering or when the index case mentioned they did not know the personal details of their risk contacts. Individuals attending the same gathering were classified as HRC in case the attack rate was high, resulting in many more registered and tested HRC compared to other index cases. This may also have impacted the KPIs obtained since September 2021.

A study evaluating the contact tracing system in Catalonia, Spain, found an increase in the proportion of index cases that had already been identified as a contact from 34% in May 2020 to 58% in November 2020.^19^ This is considerably higher than what we found for Belgium, with the proportion being at most 25% during the period May–June 2021. In addition, in our study only 42·2% of index cases were linked to at least one traced HRC, while in Catalonia 67·1% of index cases reported at least one close contact, of which 99·8% were effectively traced. A study in Portugal conducted between 1 March and 30 April 2020 compared the distribution of secondary cases between index cases that were previously identified as a close contact or returned from affected areas and those that had not been subject to contact tracing or quarantine measures before diagnosis.^20^ In line with our results, they found that ‘known’ index cases were associated to a lower number of close contacts. In contrast, they did not find a difference in the number of secondary cases or SAR between ‘known’ and ‘new’ index cases. Country-specific differences in the contact tracing setup, training of the operators, and the inquiries made, but also the absolute number of confirmed cases to process, are likely to have an impact on the effectiveness of contact tracing, though a clear comparison falls outside the scope of this study.

In addition to breaking transmission chains, contact tracing provides an important source of information regarding transmission characteristics. The observed increase in transmission from 10-19 year olds to 40-49 year olds during circulation of the Delta VoC, and similarly from 0-9 year olds to 30-39 year olds during the rise of the Omicron VoC, indicates an increased transmission potential of children for these VoCs. Although this was, to a lesser extent, also observed in March/April 2021 when the Alpha VoC was circulating, an explanation could be the increased vaccination coverage in adults compared to April 2021, when only 7% of the general population was fully vaccinated, because of the protection of vaccination against onward transmission.^21^ By the end of July 2021, 60% of the general population in Belgium was fully vaccinated, while by the end of December 2021, 75% of the general population was fully vaccinated and 38% of the adult population had received a booster vaccine.^22^ The shorter serial interval observed for ‘known’ index cases could be explained by the mandatory quarantine endorsed by the tracing, which limits their social contacts and as a result lowers their probability of transmitting the virus later in their infectious period.^17^ In addition, previous studies have reported a shorter serial interval for Omicron compared to Delta VoC, suggesting faster transmission.^23,24^ In that sense, a shorter serial interval will reduce the impact of contact tracing since the turnaround time needs to be higher which may not be achievable. This may explain the increased SAR observed in November–December 2021 when the Omicron VoC started circulating.

A previous study analyzing Belgian contact tracing data found that 8·8% of reported index cases could not be reached by telephone for an interview.^25^ In that study, self-reported data were available on the most probable place of infection for about half of the index cases. The most common place of infection was the household, followed by family/friends, work, and teenage activities including schools. In addition, the positivity ratio among unvaccinated HRC was significantly higher than among fully vaccinated HRC. Through linkage with the laboratory test database, they reported that traced HRC represented only 6% of all tests in Belgium but 24% of all new cases, indicating the effectiveness of contact tracing in case finding. In addition, they reported that only 36% of index cases were fully vaccinated by the end of September 2021, which is low compared to the vaccination coverage of 73% in the general population at that time.

In addition to operational challenges, the effectiveness of contact tracing also relies on testing policy and compliance, the cooperation of index cases, and the imposed and adopted quarantine and isolation measures. Some index cases might, willingly or unwillingly, not report all of their contacts. This could explain why the number of traced HRC observed in our study is lower than the average of 4 contacts that has been found based on social contact surveys.^26^ Even when including only those index cases that were linked to at least one traced HRC, the average was only 2·5 (IQR 1 - 3) HRC. A possible explanation is that duplicate contacts were usually linked to only one index case, in order to avoid contacting the same individuals more than once. Additionally, individual contacts that were traced as part of a collectivity are not included in the available data. Although the most important thing is to prevent onward transmission, it would be beneficial to record all individual contacts for each index case. In this way, more reliable contact and transmission networks can be reconstructed, which is important if one wants to estimate epidemiological characteristics such as the reproduction number and generation interval.^26–28^

This study has several limitations. Conversion from contact to index case could only be assessed for contacts with a known NRN, and it is not known whether a contact that does not reappear in the index dataset tested negative or was not tested at all. For about 30% of traced HRC the NRN was missing, resulting in a large part of traced HRC that could not be included in these analyses. In addition, 1.5% of traced HRC were linked more than once to the same index case, 8% were linked to an index case without a known identifier, and 3% were linked to an index case not included in the available data. From the available data, directionality of transmission cannot be assessed. It is possible that an index case was infected by one of their traced contacts, but was the first individual showing symptoms and/or to be contacted by the contact tracing. Interview data contained only aggregated information on the number of contacts in collectivities, hence this setup was not suitable for our analyses which were performed at the level of the index case. Because of specific guidelines for testing and contact tracing in collectivities such as nursing homes and schools, the available data are also not fully representative of young children and the elderly population because the majority of their contacts have been traced and followed up by local medical services. Therefore, transmission within these age groups may be underrepresented in our transmission matrices. This may also explain the low proportion of index cases aged above 85 years, as well as the low proportion of ‘known’ index cases among those aged above 65 years. An additional explanation may be the earlier vaccination of elderly, protecting them against infection.

In conclusion, the type of analyses performed in this study can be used to continuously monitor the performance and impact of contact tracing. It is difficult to fully quantify the effectiveness of contact tracing in all its dimensions without a reference period or region in which the same data are collected without informing risk contacts on their potential exposure, explaining measures to be taken, and motivating contacts to comply. However, we show that contact tracing in Belgium from September 2020 to December 2021 has been effective in reducing onward transmission. This study also shows that in times of high burden on the healthcare system, contact tracing KPIs should be interpreted with caution in light of changing testing and quarantine strategies. In addition, the data obtained by contact tracing allow to investigate transmission patterns by individual characteristics of the index case, such that contact tracing can be prioritised to individuals with a high contribution to transmission at a certain point in time.

## Data Availability

The line list data and R code will be made publicly available upon publication of the
manuscript.

## Acknowledgements

The authors are grateful for the many interesting discussions within the Controletoren Consortium including members of Hasselt University, Ghent University, KU Leuven, and University of Antwerp. Part of this project was funded by the Flemish Agency for Care and Health, which also provided the data. The funding source had no involvement in study design, analysis, nor in the decision to submit the paper for publication.

## Authors’ contributions

Cécile Kremer: conceptualisation, data curation, formal analysis, investigation, methodology, resources, software, visualisation, writing – original draft, writing – review & editing; Lander Willem: conceptualisation, data curation, investigation, methodology, resources, writing – original draft, writing – review & editing; Jorden Boone: investigation, resources, validation, writing – review & editing; Wouter Arrazola de Oñate: validation, writing – review & editing; Naïma Hammami: investigation, resources, validation, writing – review & editing; Christel Faes: conceptualistion, project administration, supervision, writing – review & editing; Niel Hens: conceptualisation, investigation, methodology, project administration, supervision, writing – original draft, writing – review & editing.

## Ethics declaration

This study has been approved by the Agency for Care and Health (GE0-1GDF2IA-WT/1GD305/20069780). It was conducted in accordance with international ethical standards (Declaration of Helsinki 1964). It was conducted in accordance to the General Data Protection Regulation (GDPR) and a data processing agreement between the Agency of Care and Health and the Universities of Antwerp and Hasselt was concluded. Participant information was coded and held securely. De-identification was performed on data content to comply with the Data Protection Regulation scope.

## Data and code availability

The line list data and R code will be made publicly available upon publication of the manuscript.

## Supplementary Figures

**Figure S1:**
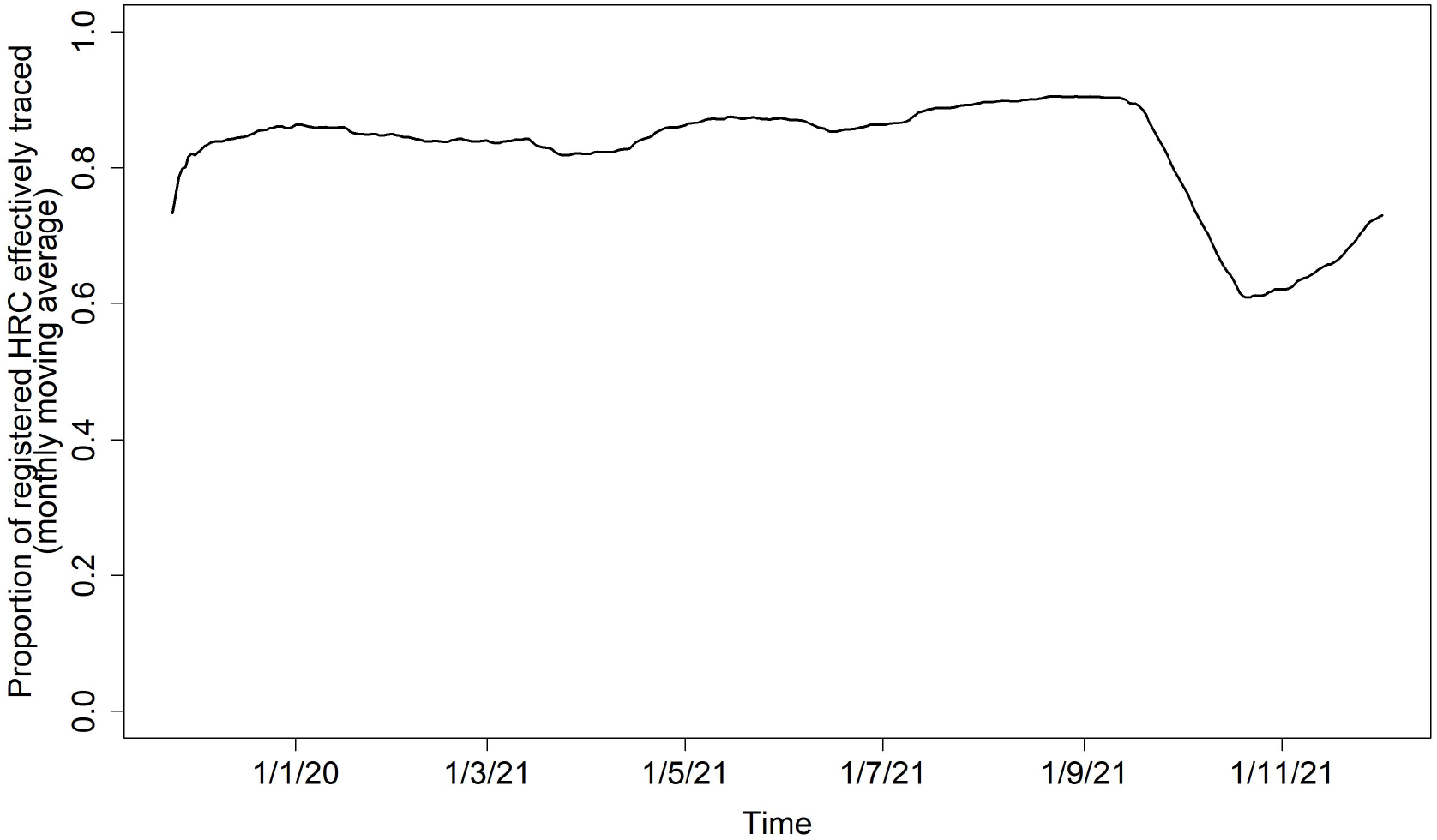
The proportion of registered high-risk contacts (HRC) that was effectively traced, over time (monthly moving average).

**Figure S2:**
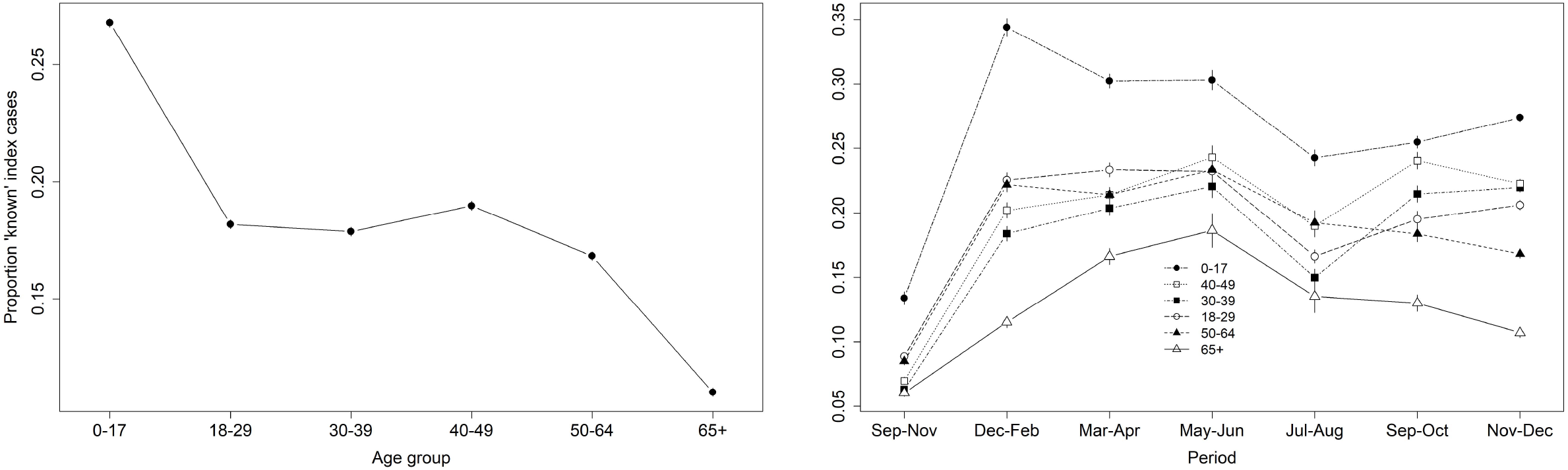
Evolution in the proportion of ‘known’ index cases by age group. Vertical bars represent the mean *±* 2*×* standard error.

**Figure S3:**
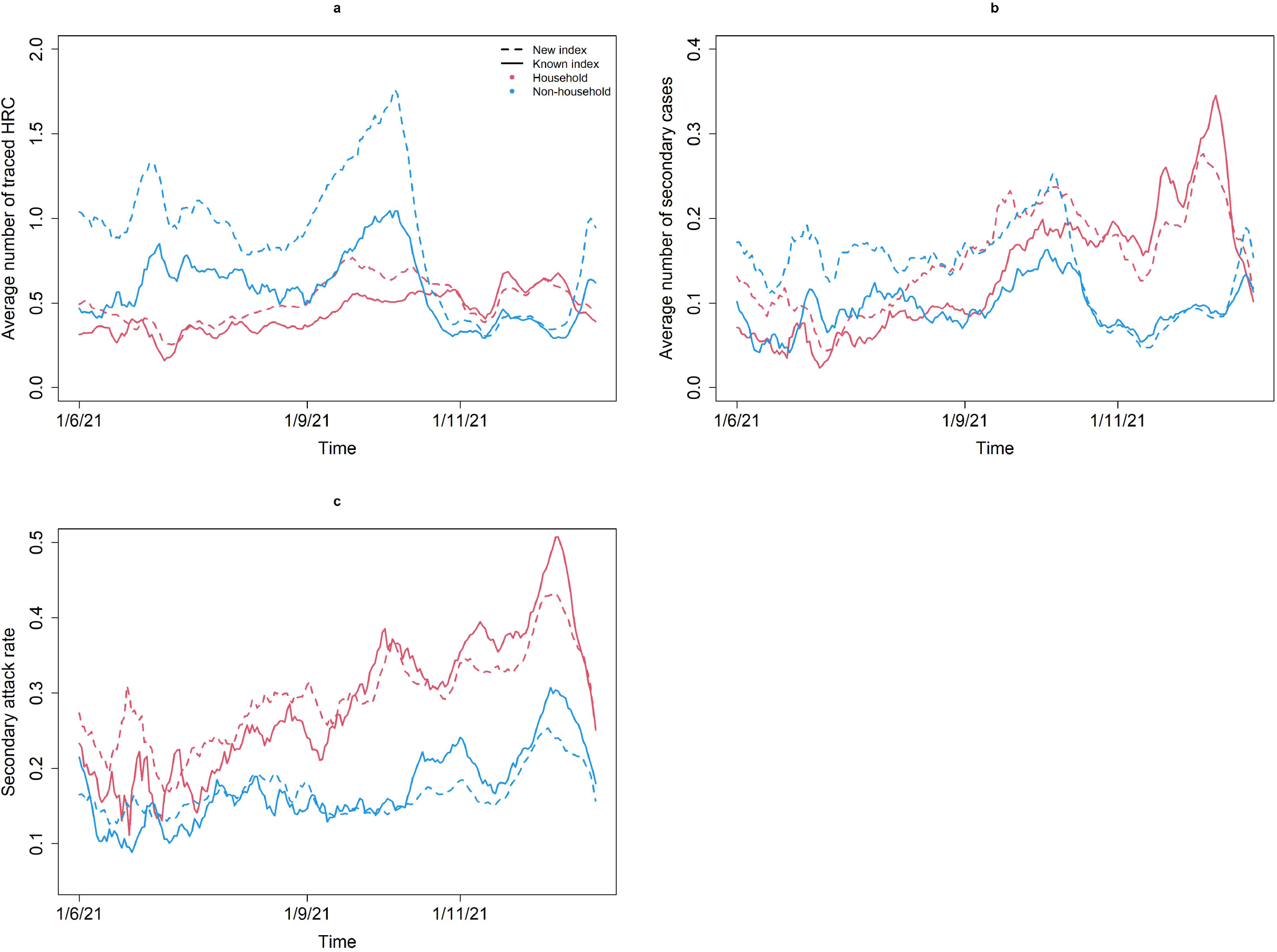
Evolution by household status in the (a) average number of traced HRC for ‘new’ and ‘known’ index cases, (b) average number of secondary cases for ‘new’ and ‘known’ index cases, and (c) secondary attack rate (SAR) among traced HRC of ‘new’ and ‘known’ index cases.

**Figure S4:**
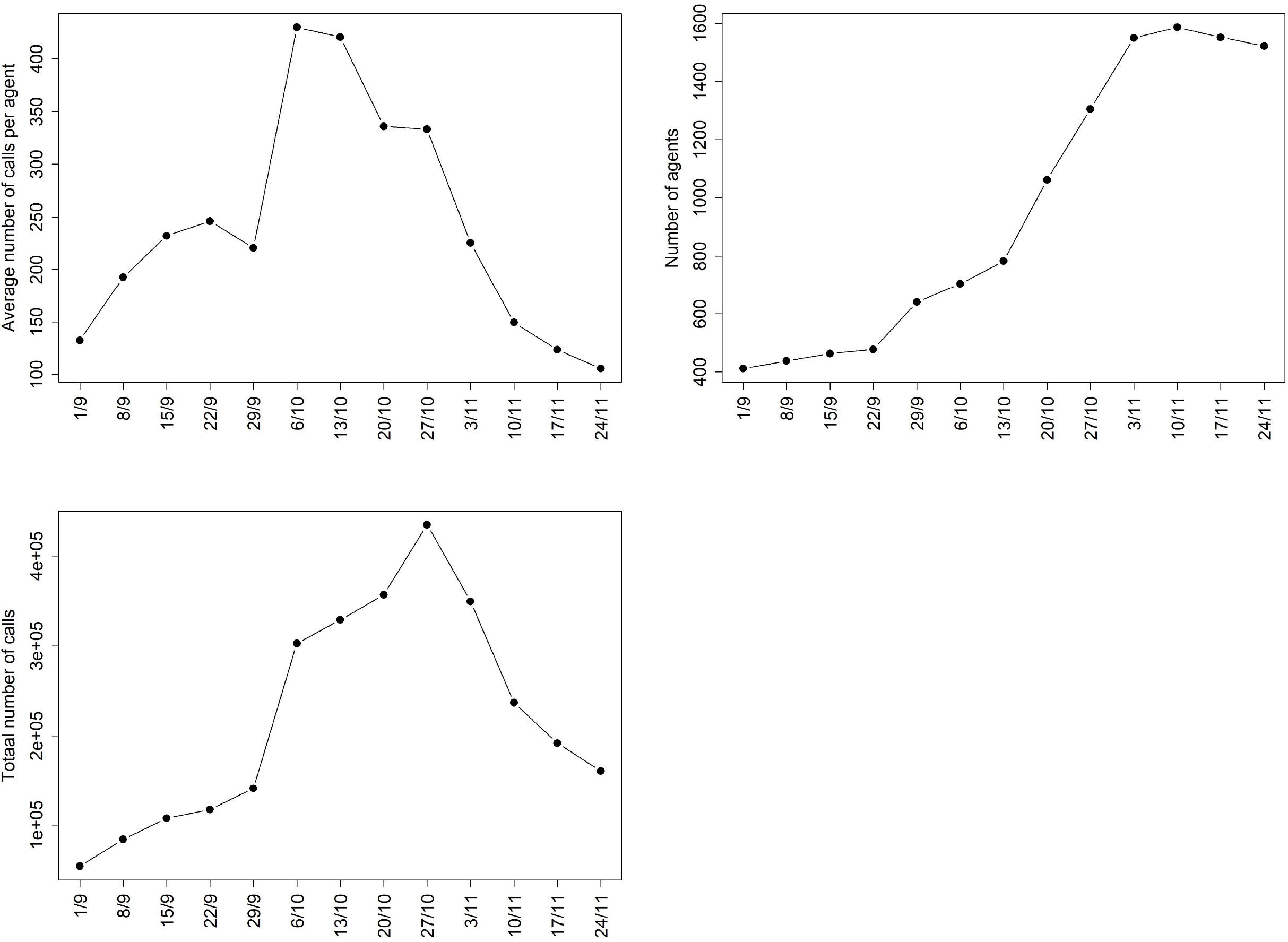
Evolution in the (a) average number of calls per agent, (b) total number of agents, and (c) total number of calls, for the period from September to November 2020.

**Figure S5:**
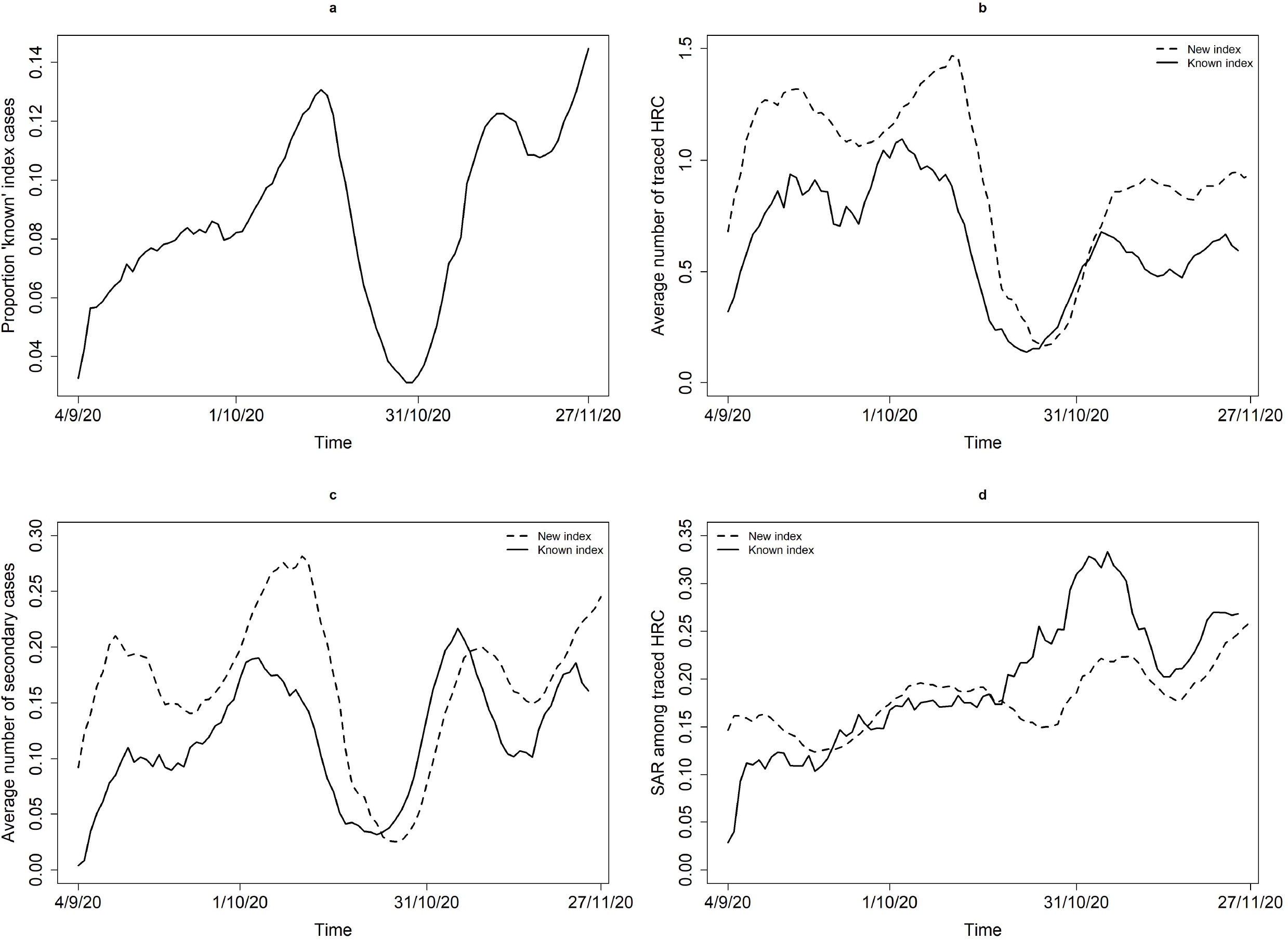
Evolution in the (a) proportion of index cases that were previously identified as a risk contact, (b) average number of traced high-risk contacts (HRC) for ‘new’ and ‘known’ index cases, (c) average number of secondary cases for ‘new’ and ‘known’ index cases, and (d) secondary attack rate (SAR) among traced HRC of ‘new’ and ‘known’ index cases, for the period from September to November 2020.

**Figure S6:**
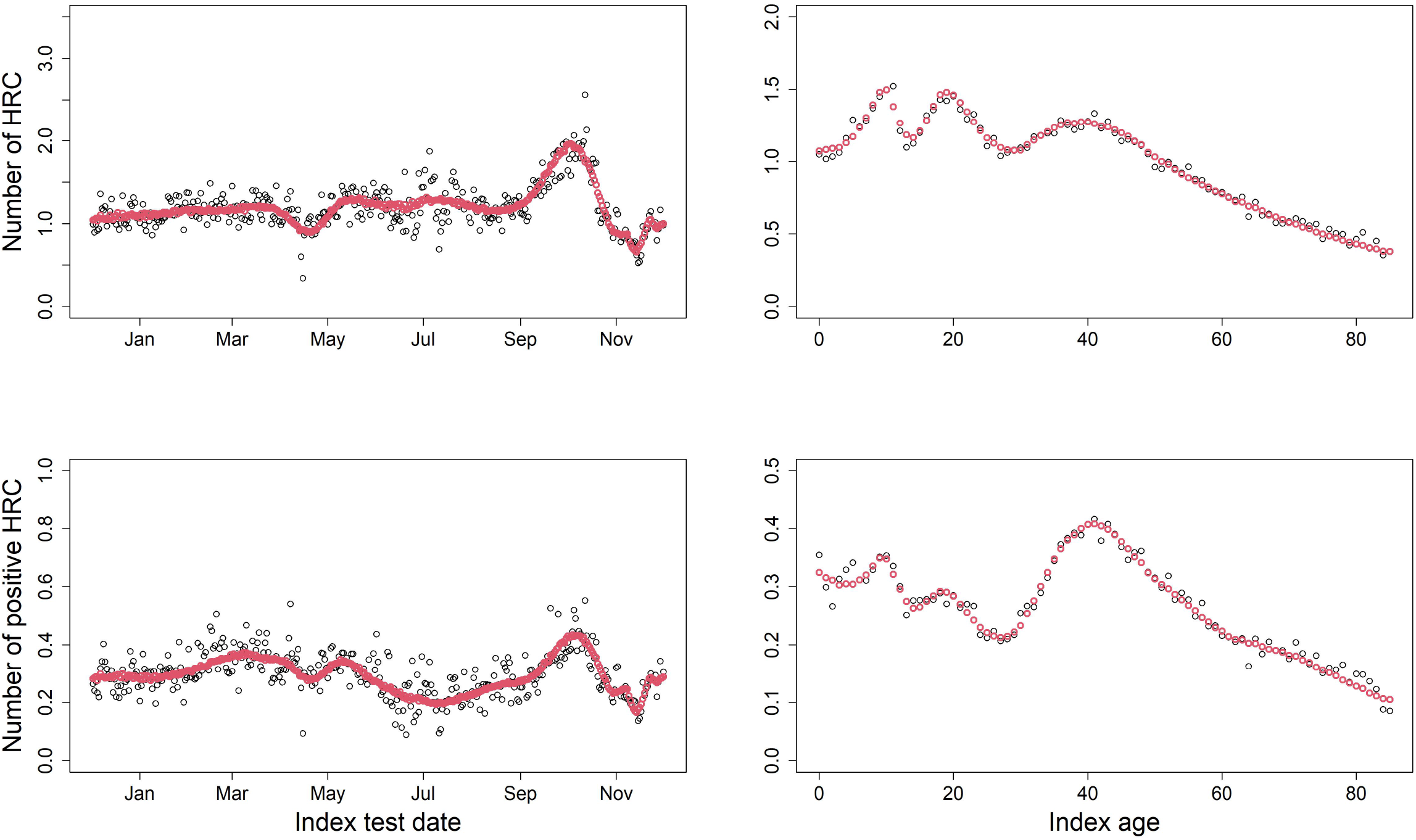
Predicted average number of (positive) HRC from negative binomial regression (red) and observed means in the testing sample (black).

